# Classifying breast cancer and fibroadenoma tissue biopsies from paraffined stain-free slides by fractal biomarkers in Fourier Ptychographic Microscopy

**DOI:** 10.1101/2023.12.22.23300439

**Authors:** Vittorio Bianco, Marika Valentino, Daniele Pirone, Lisa Miccio, Pasquale Memmolo, Valentina Brancato, Luigi Coppola, Giovanni Smaldone, Massimiliano D’Aiuto, Gennaro Mossetti, Marco Salvatore, Pietro Ferraro

## Abstract

Breast cancer is one of the most spread and monitored pathologies in high-income countries. After breast biopsy, histological tissue is stored in paraffin, sectioned and mounted. Conventional inspection of tissue slides under benchtop light microscopes involves paraffin removal and staining, typically with H&E. Then, expert pathologists are called to judge the stained slides. However, paraffin removal and staining are operator-dependent, time and resources consuming processes that can generate ambiguities due to non-uniform staining. Here we propose a novel method that can work directly on paraffined stain-free slides. The method is automatic, independent from the operator, and provides classification of different portions of the tissue image with very high accuracy. Besides, it returns a guide map to help pathologist to judge the different tissue portions based on the likelihood these can be associated to a breast cancer or fibroadenoma biomarker. We use Fourier Ptychography as a quantitative phase-contrast microscopy method, which allows accessing a very wide field of view (i.e., square millimeters) in one single image while guaranteeing at the same time high lateral resolution (i.e., 0.5 microns). This imaging method is multi-scale, since it enables looking at the big picture, i.e. the complex tissue structure and connections, with the possibility to zoom-in up to the single cell level. In order to handle this informative image content, we introduce elements of fractal geometry as a multi-scale analysis method. We show the effectiveness of fractal features in describing fibroadenoma and breast cancer from six patients with very high accuracy. The proposed method could significantly simplify the steps of tissue analysis and make it independent from the sample preparation, the skills of the lab operator and the pathologist.

## Introduction

Breast cancer represents one of the most monitored pathologies for women due to its high mortality and morbidity rate. In fact, the five-year survival rate in metastatic breast cancer is less than 30%. Recent data produced by the IARC (International Agency for Research on Cancer) report that in 185 examined countries, 2.3 million new cases (11.7%) of breast cancer were found with a mortality rate of 6.9%.^1^ Also, the incidence of breast cancer is more common in high-income countries (571/100,000) than in low-income countries (95/100,000). Breast cancer encompasses a group of diseases characterized by different biological subtypes, with a molecular profile and specific clinical-pathological characteristics.^2^ The diagnosis of breast cancer is based on clinical examination combined with imaging and confirmed by pathological assessment. The comprehensive pathological assessment of breast cancer should be performed in alignment with the World Health Organization (WHO) classification^3^ and the eighth edition of the American Joint Committee on Cancer (AJCC) Tumour, Node, Metastasis (TNM) staging system,^4^ and includes not only anatomical considerations but also crucial prognostic insights tied to tumor biology, such as tumor grade, estrogen receptor (ER), progesterone receptor (PgR), human epidermal growth factor receptor 2 (HER2), and available gene expression data.^5^

In clinical practice, the pathological assessment of breast tissue is usually performed through needle aspiration, biopsy, or surgical excision. Immunohistochemical investigation such as the classical hematoxylin and eosin (H&E), the staining with specific antibody and other useful molecular tests are used for the characterization of breast cancer. Moreover, the diagnosis of breast cancer is made by pathologists requiring time and experience. Hence, the diagnosis results do not always coincide as they depend on several factors such as previous experience and sample preparation. Currently, accuracy of diagnosis is limited to 75%.^6^ Although recent progresses on image acquisition of tissue slides at modern microscopes have brought to a strong automation, the evaluation still remains extremely complex, very time-consuming and labor-intensive. Therefore, reliable, rapid, automatic and less dependent operator methods for the breast cancer diagnosis are still challenging and far from the actual needs. Essentially, in the era of digital analysis and big data, the bottleneck in the diagnosis lies in the visual examination of such huge images that implies tiredness workload for histopathologists. One possibility is to apply straightforward approaches to obtain classifiers working on texture and morphological features by a computational analysis of standard microscope images of stained tissue slides. This would have the benefit to make steps forward in classifying benign and malignant tumors by automatic process and try to overcome the subjectivity in image analysis.^7,8^ Recently, interesting developments have been introduced in the histopathology field by digital pathology. The widespread use of slide scanning systems is mainly associated with the reduction of the costs of the scanning technology and digital storage. Nowadays, the technology advances offered by the modern microscope apparatus commonly named as Whole Slide Imaging (WSI) has opened the route to several new possibilities.^9,10^ WSI allows very fast and high-resolution acquisition of entire tissue slides thus making available images of the biopsies in digital format with typical times compatible with clinical practice.^11^ Accessing such a huge amount of data has favoured the use Artificial Intelligence (AI) by feeding Neural Networks with examples in the form of images for enhancing research and clinics in oncology. Quantitative data can be also extracted from digitized histopathology images of whole tissue slides. Such information can support and boost researchers, physicians and pathologists in the accurate analysis of a patient’s slide and can make their work faster and less cumbersome.^12-15^

Several aspects can significantly affect the quality of immunohistochemistry or, overall, the entire preparation process for tissues slides, such as storage time, oxidation, hydrolysis, tissue processing time, fixation time and type. Within this framework, the current protocols for clinical pathology are based on the inspection of stained slides, so that a pathologist is used to judge the slide based on her/his knowledge of the appearance of the morphology of a healthy tissue section when observed through the intrinsic filter brought by the stain. Hence, tissue slides preparation and staining process is crucial for an effective diagnosis. Besides, fixation in 10% neutral buffered formalin, for approximately 15 hour, and slide storage in paraffin is of great importance to preserve the tissue samples. Usually, coating or dipping coating with paraffin is provided in order to embed and seal the tissue slides to reduce oxidation. Then, for imaging the sample slides using a light microscope (LM), paraffin has to be removed in an incubator and the tissue slide has to be stained, as sketched in Fig. 1(a).^16^ However, staining the tissue using for example H&E is a process that can lead to misinterpretations since the results of the staining process strictly depend on the lab operator. Thus, an image section can appear “brighter” or “darker” depending on the laboratory where the staining has been applied, and the interpretation of the pathologist called to judge the slide can be affected as well. Similarly, algorithms for automatic image analysis and even deep learning architectures can be affected by such stain-induced ambiguities.^17^ In general, staining a sample is a process characterized by a large failure rate so that a large amount of stained slides cannot be used for histopathology purposes, e.g. due to uneven staining.^18^ Uneven staining can also be caused during the process of paraffin removal, or due to incorrect sectioning, overly dehydrated tissue, poorly infiltrated tissue, or water provoking under-staining of cytoplasmic structures.^19^ Also, the use of formalin can cause over-drying and searing of the outer edges of the tissue when the slide is excessively exposed to sunlight, thus provoking an incorrect appearance and loss of the nuclear details.^19^ Microscopy observation of the slides under a LM returns false colour images showing the stained areas with higher contrast, while the tissue inner structures that do not bind to the stain are returned with poor contrast. Such type of observation, although widely accepted and used in clinical practice, is intrinsically ambiguous due to the above-mentioned operator-dependency and also to the lack of an absolute reference to compare images having pixel values not linkable to a physical measure.^6^

**Figure 1.**
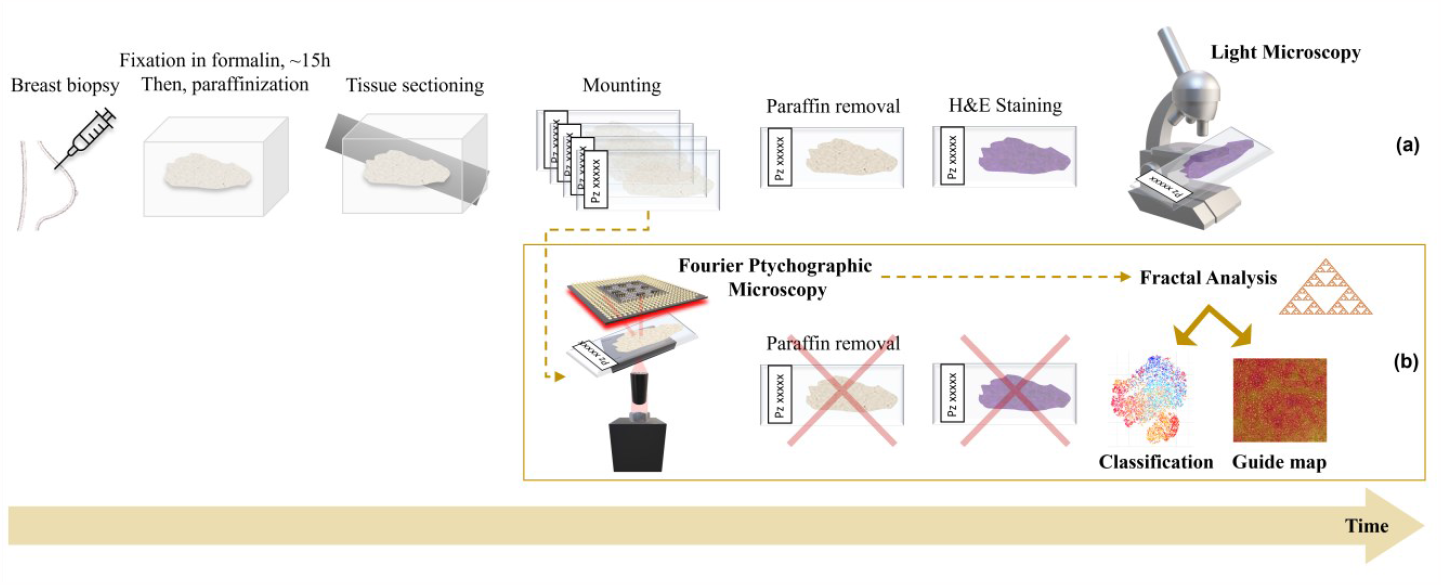
Sketch of the process of tissue slide analysis. **(a)** Conventional light microscopy analysis. **(b)** Proposed stain-free method.

With the aim to avoid the ambiguities associated with staining, label-free methods have emerged. For example, Raman spectroscopy can provide sample information about biomolecular alterations in non-destructive way but in label-free mode.^20^ Furthermore, Raman spectroscopy could be combined with machine learning (ML) to perform the spectral analysis automatic and more objective. One more way to image breast tissue was based on high-definition Fourier Transform Infrared (FT-IR) imaging to find a spectroscopy signature for cancer classification.^21^

Among these several label-free techniques, Quantitative Phase Imaging (QPI) is recently emerging as a class of methods that can image label/stain-free biological samples while providing the necessary contrast for downstream analysis, physiology and histopathology observations.^22^ QPI methods measure the optical path delay introduced by the biological sample on the light probe, which is linked to biophysical quantities. In QPI, the optical readout is the phase-contrast map. Unlike LM images of stained specimens, each pixel of a phase-contrast map is proportional to the optical thickness, i.e. the product between the physical thickness and the integral along the optical axis direction of the refractive index. Similarly, the dry mass is a quantitative parameter that can be calculated for each pixel in the image. Both these quantities depend on the local density of the specimen (e.g. cells nuclei show higher phase-contrast and dry mass than cytoplasmic compartments).^23^ This mechanism provides the contrast needed for analyzing tissue slides without ambiguities. Different QPI approaches have been proposed to inspect biological tissue slides in stain-free mode, including Digital Holography (DH),^24-26^ Fourier Ptychographic Microscopy (FPM),^27-33^ micro-optical coherence tomography,^34^ SLIM,^35^ and the above-mentioned WSI.^9,10^ Among the different QPI approaches, FPM is preferred to favourably stretch the optical constraint that limits the obtainable space bandwidth product. In FPM, phase-contrast imaging over a wide Field of View (FoV) is accessible by selecting an optical configuration adopting low magnification microscope objectives (MOs). For most of the benchtop microscopes, this choice means sacrificing the available lateral resolution. In FPM, angle diversity is introduced in the illumination pattern with the aim to enhance the resolution according to a synthetic aperture principle.^27-33^ A set of bright-field and dark-field images are captured, each one transferring a subset of spatial frequencies of the sample. In particular, the large angle light probes (dark field images) have the effect of conveying the high frequency details into the MO Numerical Aperture (NA), thus transferring them within the system bandpass cutoff. Then, a phase-retrieval process estimates the high-resolution complex amplitude from the set of low-resolution intensities. The effect is a mm^2^-cm^2^ size FoV image with submicron lateral resolution. This is ideal to investigate histopathology slides where small tissue portions are not necessarily representative of the condition of the patient undergoing biopsy and the inspection of the entire slide is sometimes necessary. FPM has been used with various coded illumination schemes^36^ for imaging cell cultures and tissue slides^27,37^ in applications ranging from biology research and drug testing to mechanobiology.^27,31,38^ Recently, deep learning methods have been employed to fasten the reconstruction process^33,39,40^ and to make FPM microscopes more robust against misalignments, thus helping the ongoing process of translating FPM to clinical practice.^32,41^

Here we use FPM to image and analyse breast tissue slides from six patients in stain-free modality. We accurately identify the tissue portions exhibiting breast cancer from the fibroadenoma areas. ML is applied by extracting meaningful features from the wrapped (i.e., modulus 2π) FPM phase-contrast maps. The features are used to train a classifier to infer the class each image patch belongs to. Then, by using a max-voting approach specifically developed for digital histopathology, the proposed method is able to provide accurate classification at the single patch level, image level, and patient’s level with increasing minimization of the classification error (i.e., on average, 21.6%, 7.7%, and 0.0%, respectively). As a result of this analysis, we provide a very accurate overall classification of the patients’ slide to furnish a first automatic indication to the pathologist. Besides, we create a heatmap of the most relevant parameter for classification, which can serve as a guide to establish the areas where the breast cancer phenotype is more or less expressed.

The stain-free process we propose is sketched in Fig. 1(b). It is important to note that in our FPM imaging of tissue slides it is not necessary to remove paraffin from the slides. Paraffin acts preserving the tissue slides. However, it can be detrimental for FPM phase imaging. Indeed, paraffin can act as a layer with large refractive index that introduces an additional optical path delay and provokes severe phase wrapping. Besides, refraction from the paraffin can change in unpredictable way the illumination vector, so that the actual illumination of the tissue portion is not consistent with the nominal illumination used in the FPM-phase retrieval algorithm. Nevertheless, we demonstrate that in FPM, paraffin does not affect the analysis when this is carried out relying on fractal biomarkers. Essentially, we show that the phase wrapping pattern obtained from FPM can be used as a fingerprint to characterize and classify the different portions of the image. In particular, with the aim of obtaining features to classify FPM images where the tissue morphology cannot be inferred, we rely on a recently developed analysis framework based on elements of fractal geometry. Fractal geometry is a branch of math particularly suitable to describe natural objects and their complexity.^42,43^ In microscopy, it has been applied to various problems, e.g. to describe the capillary system in angiography,^44^ the structure of neuron networks in LM images of brain tissue slides,^45^ to phenotype tumour cells,^46^ and in scattering-based cytometry to characterize the complexity of scattering patterns of single cells^47^ and their link to the intracellular composition and distribution of organelles, e.g. the mitochondrial network in healthy and precancerous epithelial cells.^48^ Recently, we applied the fractal analysis to wrapped holographic phase-contrast maps of marine microalgae and microplastics to define a fingerprint of microplastic items and identify them in water samples.^49^ Indeed, complexity descriptors like fractal dimension and lacunarity^44,50,51^ are particularly useful to characterize the distribution of the phase jumps within each single cell.^49^ Here we apply such elements of fractal geometry to the FPM wrapped phase-contrast images, i.e. we describe the structure of phase-jumps (or “lacunes”) at the whole image level to train the classifier. Such analysis is made possible thanks to the “multi-scale” feature of both FPM imaging and fractal geometry. We show the effectiveness of fractal descriptors to classify the stain-free digital images of breast tissue biopsies recorded by FPM without removing paraffin, as sketched in Fig. 1(b).

## Material and Methods

### FPM complex amplitude estimate

FPM^27,28^ is a non-interferometric QPI technique^29^ that provides phase images relying on phase retrieval algorithms.^30^ The FPM main feature is to generate high-resolution phase-contrast images over a large FoV. This is possible thanks to the principle of synthetic numerical aperture (PSNA),^31^ which allows to overcome the trade-off between high lateral resolution and large FoV, which is typical of conventional microscopy. In fact, in conventional microscopy, the bigger the NA the higher the resolution and the smaller the FoV and vice versa.

The FPM system is built according to the PSNA, managing the illumination source and the MO. In our configuration, the MO has low numerical aperture (NA) for ensuring the wide FoV, while the illumination source is a LEDs planar array. The system setup and acquisition working principle are sketched in Fig. 2. Sequentially turning on each LED, the object on the sample plane is probed by different LED light sources with illumination angles that depend on the LED position in the source array. The central LEDs probe the object perpendicularly to the sample plane and generate bright-field intensity images on the camera, while the outermost ones provide a beam grazing the sample at a certain angle, generating dark-field intensity images on the camera. The further away LEDs are with respect to the object, the more inclined is their beam, i.e. the greater the angle. In the frequency domain, a light beam with high illumination angle shifts the illumination NA towards high frequencies.

**Figure 2.**
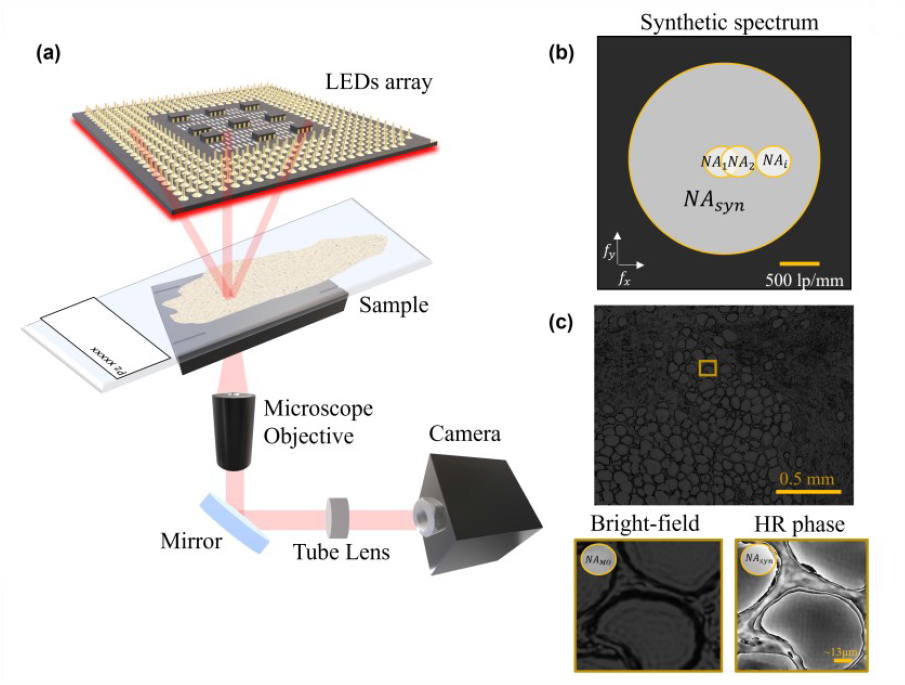
FPM acquisition scheme. **(a)** Experimental setup, where we intentionally enlarged the sketch of the sample tissue slide in the acquisition plane. **(b)** Sketch of the Fourier synthetic spectrum that shows the NA enhancement. **(c)** Top: example of bright-field image corresponding to the central LED. Bottom: zoom-in detail of the area marked by the yellow box. Bottom left: low resolution bright-field intensity. Bottom right: corresponding high resolution wrapped phase-contrast map.

Mathematically, if *O*(*r*) represents the object on the sample plane (*r* as spatial coordinate) and *e*^*j*2*πfr*^ a single LED complex field (*f* as frequency), the transmitted complex field through the object is

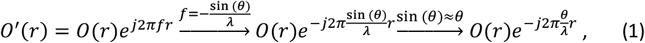

where *θ* refers to the illumination angle and *λ* to the LED wavelength. In Eq. 1, the linear correlation between the illumination angle and the frequency highlights the influence of the angle variation on the frequency values. Hence, combining properly the LED Nas in the Fourier spectrum, a bigger NA can be synthetized covering a wide frequency range (Fig. 2(b)), i.e.

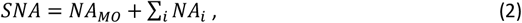

where *SNA* is the synthetic numerical aperture, *NA*_*MO*_ the numerical aperture of the MO, and *NA*_*i*_ the numerical aperture of the i-th LED.

The central LEDs contribute to image the basic structure of the object (i.e. the low spatial frequency content), while the external LEDs provide the finest details (dark field images). The captured images (per each LED) have low-resolution, and their intensities can be estimated as follows

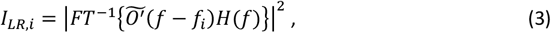

where *FT*^-1^ is the inverse Fourier Transform (FT), 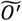 the FT of *O*^′^(*r*) and H the FT of the system impulsive response, i.e. the transfer function. An example of bright-field image of one of the breast cancer tissue slides, acquired by switching on the central LED, is reported in Fig. 2(c). In Fig. 2(c) we also show a zoom-in detail of the area marked by the yellow box. In particular, we show the enlarged detail of the low-resolution bright-field intensity and the corresponding high-resolution wrapped phase-contrast map.

The relationship between spatial domain (left side Eq. 3) and frequency domain (right side Eq. 3) is pivotal in the phase retrieval algorithm that is based on an iterative updating of the estimated complex amplitude between both domains until the convergence of the metric used is reached. After several iterations, the high-resolution complex amplitude is obtained, whose phase distribution is given by

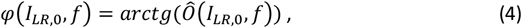

where *I*_*LR*,0_ is the initial guess of the iterative algorithm and *Ô* is the high-resolution complex field.

### FPM experimental setup

The experimental apparatus for FPM is sketched in Fig. 2. Here, we use a ×4 plan achromatic MO (Plan N, 0.1 NA, Olympus) and a 32×32 RGB LEDs array (4 mm apart), set at red wavelength (632 nm), with a bandwidth of ∼20 nm. A 400 mm tube lens converges the transmitted light beam in a charge coupled device (CCD) camera (Photometrics Evolve 512, 12-bit quantization), with 4.54 μm pixel pitch. The sample plane is 4.67 cm far from the illumination source.

An Arduino board with MATLAB^®^ codes guide the sequential illumination of 177 LEDs. The acquired images (low resolution) have a ×4.29 magnification and a size of 1460×1940. To promote the convergence of the phase retrieval algorithm and the assumption of plane wave, the images are cropped in 100×100 pixels, obtaining 266 patches. The final image (high-resolution) reaches a size of 7000×9500 pixels (where each high-resolution patch is 500×500 pixels sized). The spatial resolution of our system is demonstrated to reach 0.5 µm over a ∼3 mm^2^ FoV area.

The entire FPM process for one image takes ∼32 min by using an Intel i7-4790 CPU running @3.60 GHz and 16 GB RAM. For each patch, 7.2 s are needed to complete 60 iterations to end the phase-retrieval FP process.

### Fractal analysis of wrapped FPM maps

An example of wrapped FPM maps related to a fibroadenoma tissue slide and a breast cancer tissue slide are displayed in Figs. 3(a,c), respectively. Due to the presence of paraffin inside the imaged tissue biopsies, a dense distribution of phase jumps characterizes the wrapped FPM maps. Nevertheless, differences between the underlying tissue structures can be inferred from the wrapped FPM maps, as highlighted in the red insets in Figs. 3(a,c). In order to quantitively characterize them, an ad hoc feature set based on the fractal geometry theory was measured. At this aim, since the fractal parameters are related to a binary map consisting of full and empty areas, a zero-threshold was applied to the wrapped FPM maps. The corresponding binary FPM maps are shown in Figs. 3(b,d) for the fibroadenoma and the cancer tissue slides, respectively. Then, the binary FPM maps, made of 7000×9500 square pixels, were divided into non-overlapping 14×19 binary patches made of 500×500 square pixels, as shown by the yellow grid in Figs. 3(b,d). Moreover, as the numerical methods usually employed to compute fractal parameters work with powers of 2, each binary patch was zero-padded up to 512×512 square pixels, thus obtaining the hole patches. Instead, for each hole patch, the corresponding support patch was created by 512×512 square pixels made of 1 values. Finally, for each of the 14×19 patches, the corresponding hole patch and support patch were used to compute the 13 fractal parameters defined in Ref. ^49^, namely the fractal dimension, lacunarity index, fill ratio, regularity index, vertex density, vertex lacunarity index, vertex regularity index, fractal dimension contrast, lacunarity contrast, vertex lacunarity contrast, fractal dimension RMSE, lacunarity RMSE, and vertex lacunarity RMSE. Furthermore, for each patch, other 2 features were added, which can be related to the fractal behaviour of the wrapped FPM maps, i.e. the standard deviation and the entropy, that were computed directly from the phase values since they describe the frequency and the intensity of the phase jumps.

**Figure 3.**
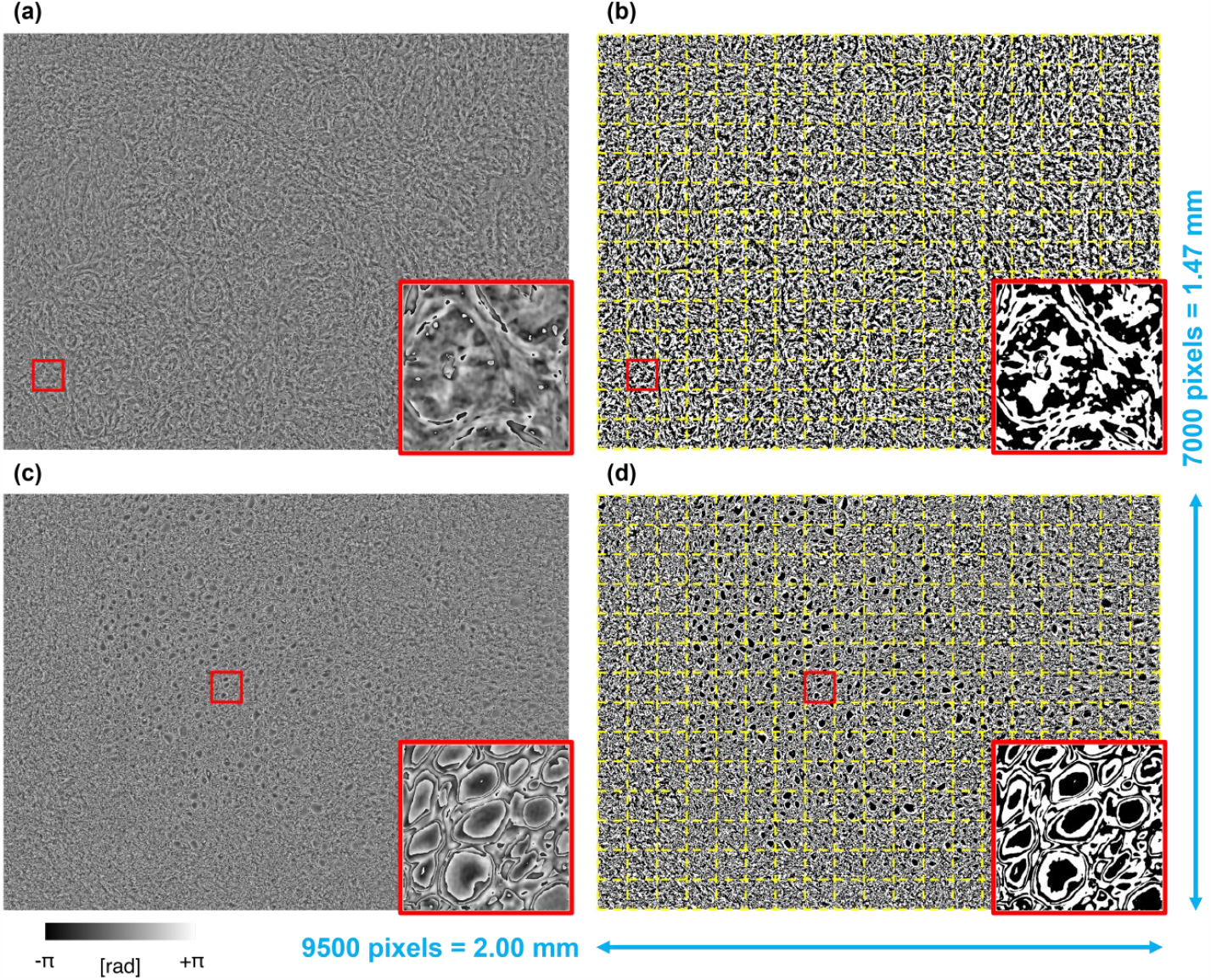
Examples of fibroadenoma (a,b) and breast cancer (c,d) FPM images. **(a,c)** Wrapped FPM maps. **(b,d)** Binary FPM maps obtained by zero-thresholding the wrapped FPM maps in (a,c), respectively, with overlapped in red the 14×19 patches (500×500 square pixels) dividing the overall 7000×9500 FOV.

## Results

### ML classification of breast tissue slides

The FPM experimental setup described in Material and Methods has been employed to image tissue biopsies taken from 6 patients, i.e. 3 patients with fibroadenoma and 3 patients with breast cancer. For each patient (i.e., for each tissue biopsy), the wrapped FPM maps of 13 different FoVs have been recorded, like those displayed in Figs. 3(a,c). According to the fractal analysis described in Material and Methods, each FoV has been divided into 266 non-overlapped patches, according to the grid sketched in Figs. 3(b,d). Finally, for each patch, 15 fractal parameters have been measured. In summary, the collected dataset is made of 15 fractal features related to the overall 20748 FPM patches, which are taken from 78 wrapped FPM maps belonging to the tissue biopsies of 6 patients (3 fibroadenoma patients and 3 breast cancer patients).

In order to inspect the collected dataset in terms of fractal features, the principal component analysis (PCA) has been implemented to reduce its dimensionality.^52^ The first three principal components are shown in Fig. 4(a), in which it can be seen that the 10374 fibroadenoma patches and the 10374 breast cancer patches shape two well defined clusters, which are quite separated each other. Furthermore, in order to perform a more detailed data inspection within the two clusters, the t-distributed stochastic neighbor embedding (t-SNE) algorithm has been exploited.^53^ Results of the t-SNE analysis are reported in Fig. 4(b), in which it can be seen that the 10374 patches belonging to the 3 fibroadenoma tissue biopsies (i.e., patients) are grouped within one single cluster (blue points). Instead, the 10374 patches belonging to the 3 breast cancer tissue biopsies (i.e., patients) form two separated clusters (red points). In particular, the left-side cluster is mainly made of patches belonging to the first and third breast cancer patient, while the right-side cluster is mainly made of patches belonging to the second and third breast cancer patient. This means that the paraffined breast cancer patches, when characterized by a fractal feature set, exhibit a greater intra-class variability with respect to the fibroadenoma patches. This is reasonable considering that the breast cancer phenotype is not homogeneously expressed over the entire FoV, rather it is localized in certain image areas. Besides, there are patches belonging to breast cancer patients that do not exhibit that phenotype and cluster in a separate region of the t-SNE diagram. Nevertheless, as well as the PCA analysis, also the t-SNE analysis confirms the good inter-class separation.

**Figure 4.**
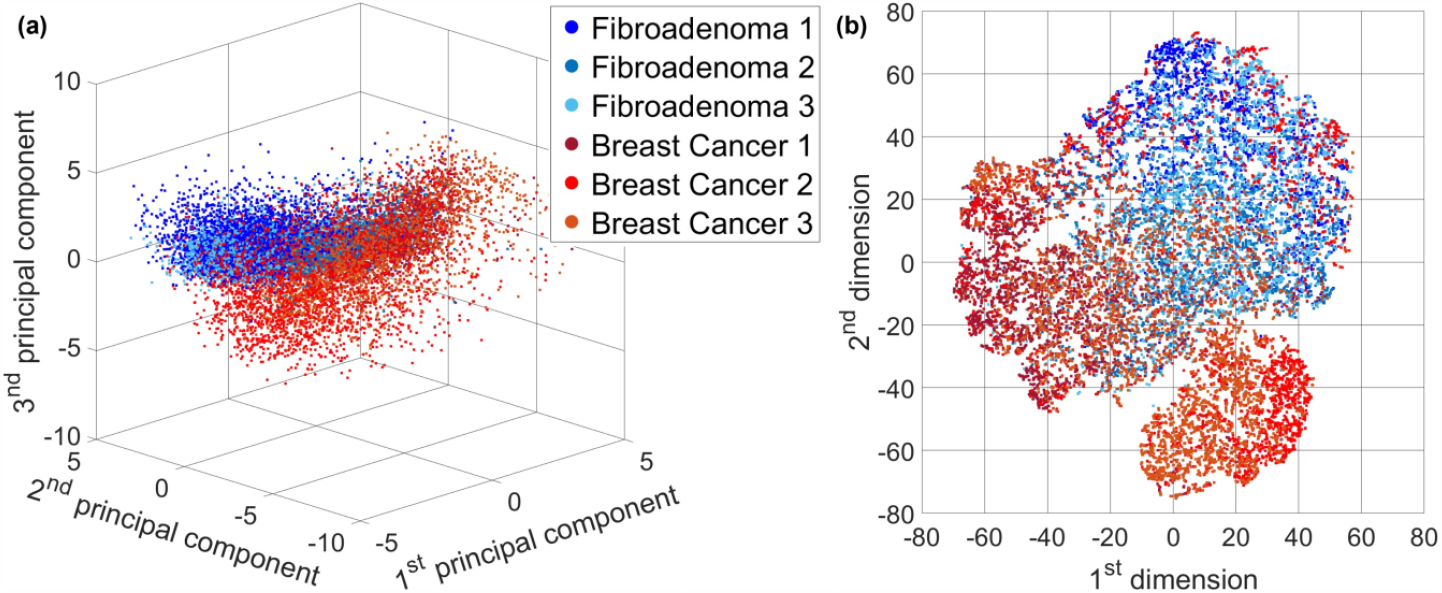
Inspection of the overall dataset by separating the contributions of the 6 tissue biopsies (3 fibroadenoma and 3 breast cancer tissue biopsies). **(a)** PCA scatter plot. **(b)** t-SNE scatter plot.

The data inspection performed in Fig. 4 suggests that the fractal feature set could be suitable to solve a classification problem for detecting a breast cancer tissue biopsy in respect to a fibroadenoma one. Moreover, the good separation between the two classes also suggests that a small training set could be enough to have a good generalization at the inference step. For this reason, the training set has been created in the worst possible condition, i.e. by using all the FPM patches of just two tissue biopsies, that are one fibroadenoma and one breast cancer patient. Moreover, to avoid any bias that could be induced by a favorable splitting of the overall dataset into a specific training set and test set, all the nine possible splits among the 6 patients have been considered. In fact, for each split, the training set is made of the 6916 FPM patches belonging to two patients (one fibroadenoma and one breast cancer patient) and the test set is made of the 13832 FPM patches belonging to the remaining 4 patients (two fibroadenoma and two breast cancer patients). For each of these nine classification problems, several ML models have been trained by using a 10-fold cross-validation.

Then, for each classification problem, it has been selected the ML model providing the best classification accuracy over the corresponding test sets, which resulted in the support vector machine (SVM) and the k-nearest neighbors (KNN).^54^ The average and standard deviation values about the resulting nine confusion matrices related to the 13832 FPM patches are summarized in Fig. 5(a), in which it can be seen that a 78.4 ± 5.0 % accuracy is reached, which is a satisfactory result considering that the training set is made of just two patients.

**Figure 5.**
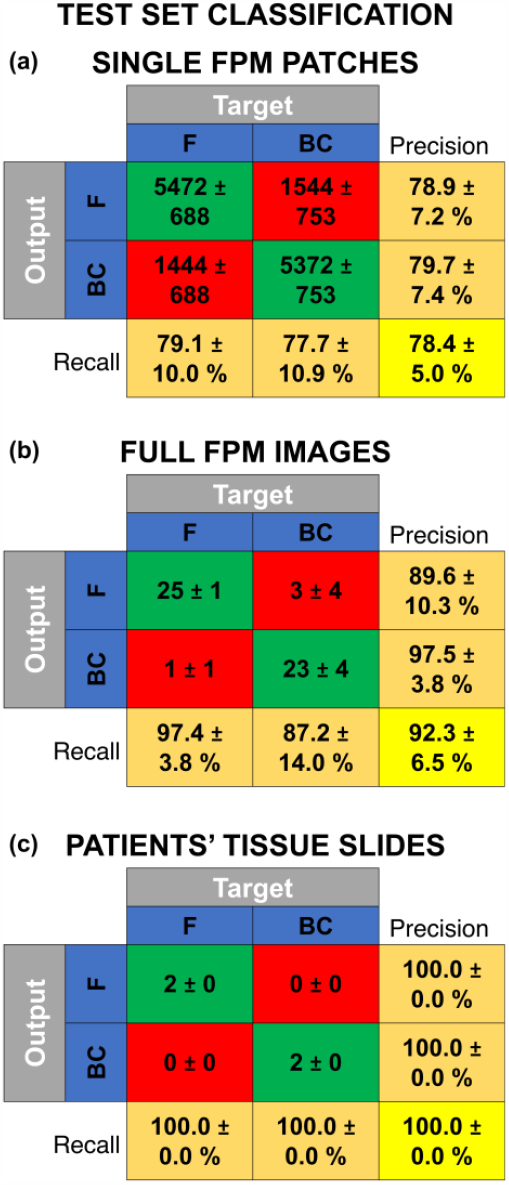
Classification between fibroadenoma (F) and breast cancer (BC) patients. **(a)** Average and standard deviation values of the nine confusion matrices obtained over the test sets made of 13832 patches belonging to 52 FoVs of 4 tissue biopsies. **(b)** Average and standard deviation values of the nine confusion matrices obtained over the test sets made of 52 FoVs belonging to the tissue biopsies of 4 patients, obtained after max-voting of the patch classes. **(c)** Average and standard deviation values of the nine confusion matrices obtained over the test sets made of 4 tissue biopsies belonging to 4 patients, obtained after max-voting of the FoV classes.

However, as discussed before, each imaged FoV is made of 266 non-overlapped FPM patches. Thus, for each FoV, 266 possible classes have been predicted by the ML classifier. This means that, in order to predict the class related to a specific FoV, a max-voting strategy can be applied,^55^ i.e. the class the imaged FOV belongs to is represented by the mode of the corresponding 266 patch classes. In this way, exploiting the intrinsic correlation between patches that belong to the same imaged FoV, the accuracy in classifying the overall FoV instead of the single patches raises up to a remarkable 92.3 ± 6.5 % within the test set made of 52 elements (see the average ± standard deviation confusion matrix in Fig. 5(b)).

In turn, 13 FOVs are imaged for each patient. Therefore, a max-voting strategy can be exploited again to combine the predicted classes of all the 13 FOVs belonging to the same slide in order to predict whether the corresponding tissue biopsy is a fibroadenoma or a breast cancer. As highlighted in Fig. 5(c), remarkably a 100.0 ± 0.0 % accuracy is reached in this classification task within the test set made of 4 patients.

### Fractal FPM heat maps as a guide for pathologists

Fractal geometry allows describing natural objects from an alternative point of view with respect to the conventional Euclidean geometry. The powerfulness of fractal geometry lies in its ability of describing patterns that are intrinsic to a certain object, thus accessing its inner complex nature. In this way, a more distinctive characterization can be extracted from the analyzed phenomenon. This is possible since fractal geometry involves a multi-scale analysis of the imaged object, meaning that it tries to describe and quantify the replication of specific patterns at different scales within the same object. For this reason, the multi-scale analysis provided by fractal geometry matches extremely well with the multi-scale imaging provided by FPM. The main advantage of FPM with respect to other imaging techniques is the large space-bandwidth product which optimizes both FoV size and lateral resolution. Hence, a large FoV can be imaged by preserving high frequency information. In this way, a biological sample can be analyzed both in its global context and in its finest details. At this scope, fractal geometry analysis can be considered the optimal solution. The fractal feature set computed from the wrapped FPM maps allows discriminating very well between fibroadenoma and breast cancer patches, as highlighted in the scatter plots of Fig. 4 and in the confusion matrix of Fig. 5(a). Moreover, accessing the wide mm^2^ FoV allows exploiting a max-voting strategy for further improving the classification performance (see Figs. 5(b,c)), since each FPM map can be divided into hundreds of patches without losing the possibility of performing a fractal characterization at the single-patch level.

In addition to the remarkable classification performance, the combination between fractal geometry and FPM also allows to generate a further source of information that could be meaningful as a guide for more in-depth studies by pathologists, i.e. fractal heat maps. In particular, among the several fractal features, the lacunarity index has been often exploited due to its higher correlation with biological phenomena.^42^ For example, lacunarity has been employed in the magnetic resonance imaging for distinguishing benign and malignant breast cancer^56^ or for differentiating the grades of glioma.^57^ It has been also used in other microscopy imaging techniques as prognostic indicator of clinical outcome in early breast cancer,^58^ for the diagnosis^59^ and the identification of the severity level of prostate cancer,^60,61^ or for the detection of the Alzheimer’s disease.^62^ Actually, the lacunarity index measures the distribution of the hole sizes within a certain structure.^51^

In the proposed study, the lacunarity index characterizes the hole maps obtained from the wrapped FPM maps, as shown in Fig. 3. In particular, for each patch in Figs. 3(b,d), a lacunarity index has been computed, as displayed in Figs. S1(a,b), respectively. In Fig. S1, each 500×500 patch takes a homogeneous value, that is the corresponding lacunarity index. Hence, images in Fig. S1 can be defined as the lacunarity heat maps. As the wrapped FPM maps have a high density of phase jumps due to the presence of paraffin within the tissue slides (see Figs. 3(a,c)), it is difficult to correlate them to a specific biological structure by means of a visual inspection. Instead, for the sole purpose of a visual analysis performed by the pathologist, the low-resolution bright-field maps (1400×1900 square pixels) can be exploited, as displayed in Figs. 6(a,c), corresponding to the wrapped FPM maps in Figs. 3(a,c), respectively. Furthermore, to help the visual analysis of the pathologist, the lacunarity heat maps computed through the quantitative fractal characterization of the wrapped FPM maps can be exploited. In particular, the 7000×9500 high-resolution lacunarity heat maps displayed in Figs. S1(a,b) can be resized to 1400×1900 square pixels in order to fit the size of the low-resolution bright-field map shown in Figs. 6(a,c), thus finally overlapping them in Figs. 6(b,d), respectively.

**Figure 6.**
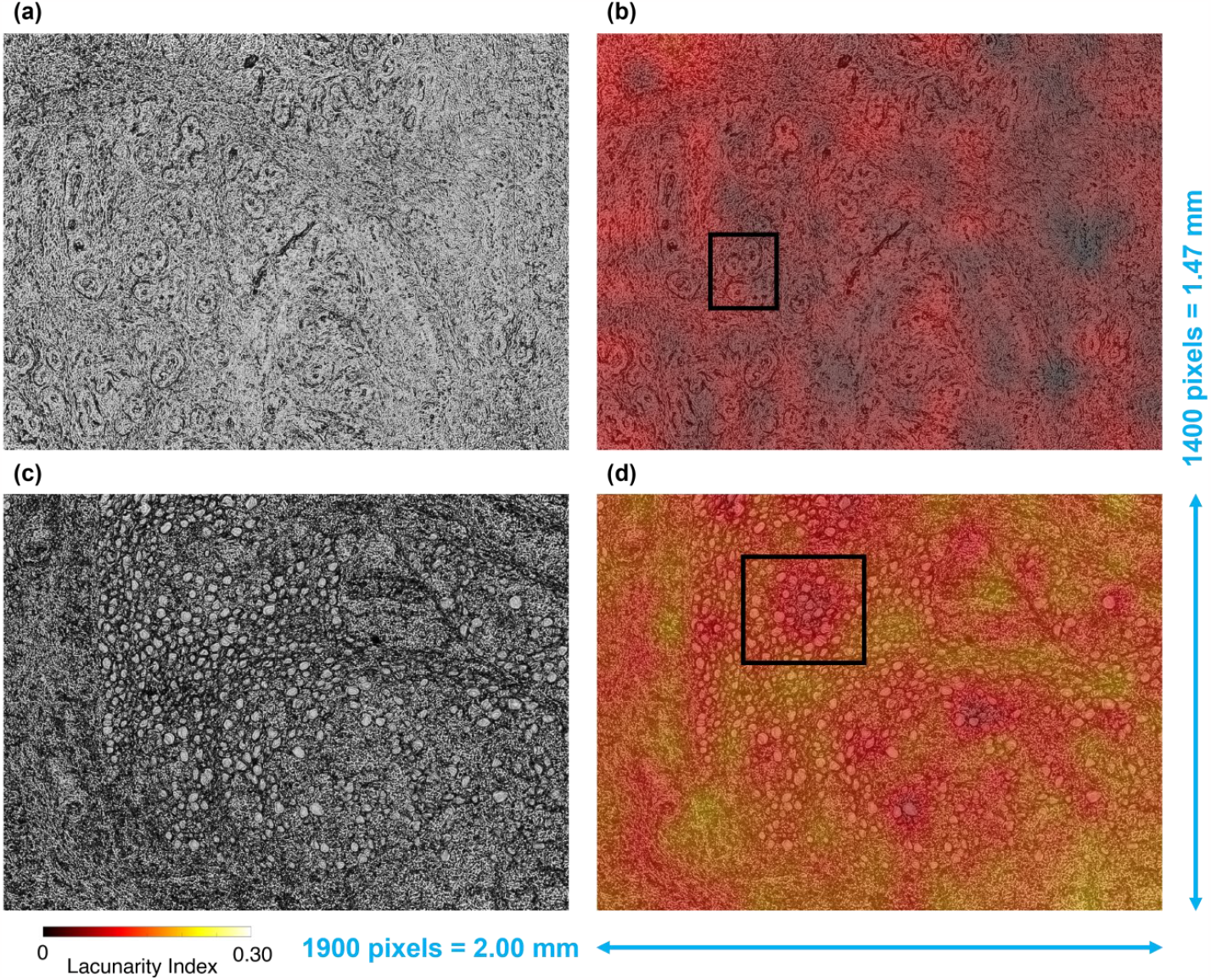
Visual inspection of the fibroadenoma (a,b) and breast cancer (c,d) tissue slides corresponding to the wrapped FPM maps in Figs. 2(a,b) and Figs. 2(c,d), respectively. **(a,c)** Low-resolution bright-field map. **(b,d)** Lacunarity heat maps of Figs. S1(a,b), down-sampled and overlapped to the low-resolution bright-field maps in (a,c), respectively. Boxes in (b,d) highlight glandular structures.

It is worth noting that the fibroadenoma tissue slide exhibits lower lacunarity indices than a breast cancer tissue slide, as respectively shown in Figs. 6(b,d). According to the definition of lacunarity index,^49^ this means that the fibroadenoma tissue slide is more lacunar than the breast cancer tissue slide. This property can be related to the different inner structures forming the two kinds of tissues, that can be seen in the low-resolution bright-field maps in Figs. 6 (a,c). Small and rather spaced ductal-derived glandular structures are observed, in a fibrous stroma in Fig. 6 (a,b), and the glandular density appears reduced. The heatmap in Fig. 6(b) highlights glandular spaced structures (see the black box). In Figs. 6(c,d), numerous ductal-derived glandular elements are observed tightly packed and without evident stroma. Glandular density appears high. Remarkably, the heatmap in Fig. 6(d) highlights numerous glandular structures bundled together (see black box).

## Discussion and Conclusions

Digital pathology analysis of breast tissue slides is widespread and can furnish a valuable help to guide pathologists called to judge heterogeneous morphologies. Extraction of features from stained histology slides obtained through WSI has emerged as a pivotal technique in pathomic studies. The primary objective is to quantitatively characterize cells and tissues derived from examined samples. Notably, some pathomic studies delve into the fractal dimension analysis to extract features to analyse WSIs of various cancer types. For instance, Lee et al. developed a computer-aided technique for the automated grading of prostatic carcinoma leveraging the application of fractal dimension for analysing pathological image texture of prostatic carcinoma WSIs. The extracted fractal dimension-based features were able to classify pathological prostate images into four classes within the Gleason grading system.^63^ Furthermore, Da Silva et al. embraced fractal dimension analysis as a computationally accessible approach to enhance the histopathological diagnosis of breast cancer. Their investigation revealed that fractal dimension-based features extracted from stained WSI demonstrated remarkable capabilities in distinguishing breast carcinomas from normal tissue and benign breast alterations. These findings underscore the significance of fractal dimension analysis as a powerful tool in advancing our understanding and diagnostic capabilities in histopathological studies.^64^

It is important to note that the histopathological analysis of breast cancer that we used as test case of our Digital Pathology approach can be critical due to its intrinsic complexity. Indeed, cell density and morphology alone cannot be sufficient for the differential diagnosis between a malignant and a benign lesion, as special cases such as florid adenosis or sclerosing adenosis could create outputs that may be associated with false-positive subjects. In addition, there are cases such as tubular breast carcinoma in which the tissue is well differentiated and only a detailed diagnostic study can make a correct diagnosis.^65^

However, independently form the specific test case in our research, we established a novel analysis framework that allowed us to analyze directly the paraffined unstained breast slides, in order to get rid of ambiguities that can be provoked by the paraffin removal and staining processes as occurs in conventional breast cancer diagnosis. We used two multi-scale methodologies for image acquisition and analysis, respectively. Fractal biomarkers, in particular the lacunarity index, well describe the FPM phase-contrast maps and allow classifying data from the single image portion level up to the patient level. In summary, the main contributions in this work are as follows:

1. We proposed an automatic method for classification of breast cancer and fibroadenoma based on the novel FPM technique applied to unstained tissue slides;
2. We introduced the fractal patterns analysis of wrapped FPM phase-contrast maps.
3. Histopathological image recognition, patch and patient classification is demonstrated by avoiding removing the paraffin layer;
4. Max-voting among different portions of the same image is demonstrated to enforce image classification. Besides, max-voting among different images from the same patient’s slide allowed very accurate classification. In particular, a 100% accuracy was obtained among the test tissue slides of 4 patients after training a ML model with the tissue slides of other 2 patients. The robustness of this method would allow to judge even using a reduced set of FPM images for the same patient.
5. The most important fractal parameter, i.e. the lacunarity index, can serve to create guide maps for pathologists.

We believe that our approach is highly innovative and potentially usable in the future to support the pathologist’s activities. In principle, the proposed strategy could be extended to other types of tissues. Therefore, next studies will be focused on the even more borderline cases mentioned above about breast cancer and other types of tissues and pathologies will be tested.

## Data Availability

All data produced in the present study are available upon reasonable request to the authors

## Acknowledgments

This work was supported by project POR CIRO (Campania Imaging for Research in Oncology) funded by Regione Campania (Italy).

This work was partially supported by the Italian Ministry of Health (“Ricerca Corrente” project).

## Data Availability

All data produced in the present study are available upon reasonable request to the authors.

## Ethics Approval and Consent to Participate

The study was approved by the Ethics Committee of IRCCS Pascale (Naples, Italy) with reference number 3/19 approved on 29 May 2019. All methods were performed in compliance with standard operating procedures and in accordance with the Declaration of Helsinki and each patient participated in the study by signing written informed consent.

## References

1. Bray F, Ferlay J, Soerjomataram I, Siegel RL, Torre LA, Jemal A. Global cancer statistics 2018: GLOBOCAN estimates of incidence and mortality worldwide for 36 cancers in 185 countries. CA Cancer J. Clin. 2018;68(6):394–424. 10.3322/caac.21492

2. Kashyap D, Pal D, Sharma R et al. Global increase in breast cancer incidence: risk factors and preventive measures. Biomed. Res. Int. 2022;2022:9605439. 10.1155/2022/9605439

3. Tan PH, Ellis I, Allison K et al. The 2019 World Health Organization classification of tumours of the breast. Histopathology 2020;77:181–185. 10.1111/his.14091

4. Amin MB, Greene FL, Edge SB et al. The eighth edition AJCC cancer staging manual: continuing to build a bridge from a population-based to a more “personalized” approach to cancer staging. CA Cancer J. Clin. 2017;67(2):93–99. 10.3322/caac.21388

5. Li J, Chen Z, Su K, Zeng J. Clinicopathological classification and traditional prognostic indicators of breast cancer. Int. J. Clin. Exp. Pathol. 2015;8(7):8500.

6. Elmore JG, Longton GM, Carney PA et al. Diagnostic concordance among pathologists interpreting breast biopsy specimens. JAMA 2015;313(11):1122–1132. 10.1001/jama.2015.1405

7. Hao Y, Qiao S, Zhang L et al. Breast cancer histopathological images recognition based on low dimensional three-channel features. Front. Oncol. 2021;11:657560. 10.3389/fonc.2021.657560

8. Wei M, Du Y, Wu X et al. A benign and malignant breast tumor classification method via efficiently combining texture and morphological features on ultrasound images. Comput. Math. Methods Med. 2020;2020:5894010. 10.1155/2020/5894010

9. Khened M, Kori A, Rajkumar H, Krishnamurthi G, Srinivasan B. A generalized deep learning framework for whole-slide image segmentation and analysis. Sci. Rep. 2021;11(1):11579. 10.1038/s41598-021-90444-8

10. Kumar N, Gupta R, Gupta S. Whole slide imaging (WSI) in pathology: current perspectives and future directions. J. Digit. Imaging 2020;33(4):1034–1040. 10.1007/s10278-020-00351-z

11. Aswathy MA, Jagannath M. Detection of breast cancer on digital histopathology images: Present status and future possibilities. Inform. Med. Unlocked 2017;8:74–79. 10.1016/j.imu.2016.11.001

12. Swillens JE, Nagtegaal ID, Engels S, Lugli A, Hermens RP, van der Laak JA. Pathologists’ first opinions on barriers and facilitators of computational pathology adoption in oncological pathology: an international study. Oncogene 2023;42(38):2816–2827. 10.1038/s41388-023-02797-1

13. Shmatko A, Ghaffari Laleh N, Gerstung M, Kather JN Artificial intelligence in histopathology: enhancing cancer research and clinical oncology. Nature Cancer 2022;3(9):1026–1038. 10.1038/s43018-022-00436-4

14. Kann BH, Hosny A, Aerts HJ. Artificial intelligence for clinical oncology. Cancer Cell 2021;39(7):916–927. 10.1016/j.ccell.2021.04.002

15. Verdicchio M, Brancato V, Cavaliere C, Isgrò F, Salvatore M, Aiello M. A pathomic approach for tumor-infiltrating lymphocytes classification on breast cancer digital pathology images. Heliyon 2023;9:e14371. 10.1016/j.heliyon.2023.e14371

16. Economou M, Schöni L, Hammer C, Galván JA, Mueller DE, Zlobec I. Proper paraffin slide storage is crucial for translational research projects involving immunohistochemistry stains. Clin. Transl. Med. 2014;3(1):1–3. 10.1186/2001-1326-3-4

17. Rivenson Y, Wang H, Wei Z et al. Virtual histological staining of unlabelled tissue-autofluorescence images via deep learning. Nat. Biomed. Eng. 2019;3(6):466–477. 10.1038/s41551-019-0362-y

18. National Diagnostics. Accessed 20 November 2023. https://www.nationaldiagnostics.com; 2023.

19. Leica Biosystems. Accessed 21 December 2023. https://www.leicabiosystems.com/it-it/knowledge-pathway/he-basics-part-4-troubleshooting-he/; 2023.

20. Ma D, Shang L, Tang J, Bao Y, Fu J, Yin J. Classifying breast cancer tissue by Raman spectroscopy with one-dimensional convolutional neural network. Spectrochim. Acta A Mol. Biomol. Spectrosc. 2021;256:119732. 10.1016/j.saa.2021.119732

21. Mittal S, Wrobel TP, Walsh M, Kajdacsy-Balla A, Bhargava R. Breast cancer histopathology using infrared spectroscopic imaging: The impact of instrumental configurations. Clinical Spectroscopy 2021;3:100006. 10.1016/j.clispe.2021.100006

22. Fanous MJ, Pillar N, Ozcan A. Digital staining facilitates biomedical microscopy. Front. Bioinform. 2023;3:1243663. 10.3389%2Ffbinf.2023.1243663

23. Bianco V, D’Agostino M, Pirone D et al. Label-Free Intracellular Multi-Specificity in Yeast Cells by Phase-Contrast Tomographic Flow Cytometry. Small Methods 2023;7:2300447. 10.1002/smtd.202300447

24. Wu D, Luo J, Huang G et al. Imaging biological tissue with high-throughput single-pixel compressive holography. Nat. Commun. 2021;12(1):4712. 10.1038/s41467-021-24990-0

25. Luo W, Greenbaum A, Zhang Y, Ozcan A. Synthetic aperture-based on-chip microscopy. Light: Sci. Appl. 2015;4(3):e261–e261. 10.1038/lsa.2015.34

26. Thomas L, Sheeja MK. Digital holographic technique based breast cancer detection using transfer learning method. J. Biophoton. 2023;16(8):e202200359. 10.1002/jbio.202200359

27. Wang T, Jiang S, Song P et al. Optical ptychography for biomedical imaging: recent progress and future directions. Biomed. Opt. Express 2023;14(2):489–532. 10.1364/BOE.480685

28. Zheng G, Shen C, Jiang S, Song P, Yang C. Concept, implementations and applications of Fourier ptychography. Nat. Rev. Phys. 2021;3(3):207–223. 10.1038/s42254-021-00280-y

29. Nguyen TL, Pradeep S, Judson-Torres RL, Reed J, Teitell MA, Zangle TA. Quantitative phase imaging: recent advances and expanding potential in biomedicine. ACS Nano 2022;16(8):11516–11544. 10.1021/acsnano.1c11507

30. Yeh LH, Dong J, Zhong J et al. Experimental robustness of Fourier ptychography phase retrieval algorithms. Opt. Express 2015;23(26):33214–33240. 10.1364/OE.23.033214

31. Ou X, Horstmeyer R, Zheng G, Yang C. High numerical aperture Fourier ptychography: principle, implementation and characterization. Opt. Express 2015;23(3):3472–3491. 10.1364/OE.23.003472

32. Bianco V, Mandracchia B, Běhal J, Barone D, Memmolo P, Ferraro P. Miscalibration-tolerant Fourier ptychography. IEEE J. Sel. Top. Quantum Electron. 2020;27(4):1–17. 10.1109/JSTQE.2020.3025717

33. Bianco V, Priscoli MD, Pirone D et al. Deep learning-based, misalignment resilient, real-time Fourier Ptychographic Microscopy reconstruction of biological tissue slides. IEEE J. Sel. Top. Quantum Electron. 2022;28(4):1–10. 10.1109/JSTQE.2022.3154236

34. Tang H, Liu X, Chen S et al. Estimation of refractive index for biological tissue using microoptical coherence tomography. IEEE. Trans. Biomed. Eng. 2018;66(6):1803–1809. 10.1109/TBME.2018.2885844

35. Majeed H, Kandel ME, Han K et al. Breast cancer diagnosis using spatial light interference microscopy. J. Biomed. Opt. 2015;20(11):111210–111210. 10.1117/1.JBO.20.11.111210

36. Sun J, Zuo C, Zhang J, Fan Y, Chen Q. High-speed Fourier ptychographic microscopy based on programmable annular illuminations. Sci. Rep. 2018;8(1):7669. 10.1038/s41598-018-25797-8

37. Valentino M, Bianco V, Miccio L et al. Beyond conventional microscopy: Observing kidney tissues by means of fourier ptychography. Front. Physiol. 2023;14:206. https://www.frontiersin.org/articles/10.3389/fphys.2023.1120099/ (10.5281/z enodo.7418567)

38. Pirone D, Bianco V, Valentino M. Fourier ptychographic microscope allows multi-scale monitoring of cells layout onto micropatterned substrates. Opt. Lasers Eng. 2022;156:107103. 10.1016/j.optlaseng.2022.107103

39. Nguyen T, Xue Y, Li Y, Tian L, Nehmetallah G. Deep learning approach for Fourier ptychography microscopy. Opt. Express 2018;26(20):26470–26484. 10.1364/OE.26.026470

40. Zhang J, Xu T, Shen Z, Qiao Y, Zhang Y. Fourier ptychographic microscopy reconstruction with multiscale deep residual network. Opt. Express 2019;27(6):8612–8625. 10.1364/OE.27.008612

41. Eckert R, Phillips ZF, Waller L. Efficient illumination angle self-calibration in Fourier ptychography. Appl. Opt. 2018;57(19):5434–5442. 10.1364/AO.57.005434

42. Losa, GA, Merlini D, Nonnenmacher TF, Weibel ER. Fractals in Biology and Medicine. Springer; 2005.

43. Franceschetti G, Riccio D. Scattering, Natural Surfaces, and Fractals. Elsevier; 2007.

44. Landini G, Murray PI, Misson GP. Local connected fractal dimensions and lacunarity analyses of 60 degrees fluorescein angiograms. Invest. Ophthalmol. Vis. Sci. 1995;36(13):2749–2755.

45. Krohn S, Froeling M, Leemans A. et al. Evaluation of the 3D fractal dimension as a marker of structural brain complexity in multiple-acquisition MRI. Hum. Brain Mapp. 2019;40(11):3299–3320. 10.1002/hbm.24599

46. Klein K, Maier T, Hirschfeld-Warneken VC, Spatz JP. Marker-free phenotyping of tumor cells by fractal analysis of reflection interference contrast microscopy images. Nano Lett. 2013;13(11):5474–5479. 10.1021/nl4030402

47. Ding H, Wang Z, Nguyen F, Boppart SA, Popescu G. Fourier transform light scattering of inhomogeneous and dynamic structures. Phys. Rev. Lett. 2008;101(23):238102. 10.1103/PhysRevLett.101.238102

48. Xylas J, Quinn KP, Hunter M, Georgakoudi I. Improved Fourier-based characterization of intracellular fractal features. Opt. Express 2012;20(21):23442–23455. 10.1364/OE.20.023442

49. Bianco V, Pirone D, Memmolo P, Merola F, Ferraro P. Identification of microplastics based on the fractal properties of their holographic fingerprint. ACS Photonics 2021;8(7):2148–2157. 10.1021/acsphotonics.1c00591

50. Karperien AL, Jelinek HF. Fractal, multifractal, and lacunarity analysis of microglia in tissue engineering. Front. Bioeng. Biotechnol. 2015;3:51. 10.3389/fbioe.2015.00051

51. Plotnick RE, Gardner RH, Hargrove WW, Prestegaard K, Perlmutter M. Lacunarity analysis: a general technique for the analysis of spatial patterns. Phys. Rev. E 1996;53(5):5461. 10.1103/PhysRevE.53.5461

52. Greenacre M, Groenen PJ, Hastie T, d’Enza AI, Markos A, Tuzhilina E. Principal component analysis. Nat. Rev. Methods Primers 2022;2(1):100. 10.1038/s43586-022-00184-w

53. Van der Maaten L, Hinton G. Visualizing data using t-SNE. J. Mach. Learn. Res. 2008;9(11):2579–2605.

54. Singh A, Thakur N, Sharma A. A review of supervised machine learning algorithms. In 2016 3rd International Conference on Computing for Sustainable Global Development (INDIACom), pp. 1310–1315.

55. Pirone D, Montella A, Sirico D et al. Phenotyping neuroblastoma cells through intelligent scrutiny of stain-free biomarkers in holographic flow cytometry. APL Bioeng. 2023;7(3):036118. 10.1063/5.0159399

56. Soares F, Janela F, Pereira M, Seabra J, Freire MM. 3D lacunarity in multifractal analysis of breast tumor lesions in dynamic contrast-enhanced magnetic resonance imaging. IEEE Trans. Image Process. 2013;22(11):4422–4435. 10.1109/TIP.2013.2273669

57. Smitha KA, Gupta AK, Jayasree RS. Fractal analysis: fractal dimension and lacunarity from MR images for differentiating the grades of glioma. Phys. Med. Biol. 2015;60(17):6937. 10.1088/0031-9155/60/17/6937

58. Pribic J, Vasiljevic J, Kanjer K et al. Fractal dimension and lacunarity of tumor microscopic images as prognostic indicators of clinical outcome in early breast cancer. Biomark. Med. 2015;9(12):1279–1277. 10.2217/bmm.15.102

59. Neves LA, Nascimento MZ, Oliveira DLL et al. Multi-scale lacunarity as an alternative to quantify and diagnose the behavior of prostate cancer. Expert Syst. Appl. 2014;41(11):5017–5029. 10.1016/j.eswa.2014.02.048

60. Waliszewski, P. The quantitative criteria based on the fractal dimensions, entropy, and lacunarity for the spatial distribution of cancer cell nuclei enable identification of low or high aggressive prostate carcinomas. Front. Physiol. 2016;7:34. 10.3389/fphys.2016.00034

61. Aralica G, Šarec Ivelj M, Pačić A et al. Prognostic Significance of Lacunarity in Preoperative Biopsy of Colorectal Cancer. Pathol. Oncol. Res. 2020;26:2567–2576. 10.1007/s12253-020-00851-x

62. Bordescu D, Paun MA, Paun VA, Paun VP. Fractal analysis of Neuroimagistic. Lacunarity degree, a precious indicator in the detection of Alzheimer’s disease. Univ. Politeh. Buchar. Sci. Bull. Ser. A Appl. Math. Phys. 2018;80:309–320.

63. Lee CH, Huang PW. Classification for pathological prostate images based on fractal analysis. In 2008 Congress on Image and Signal Processing (Vol. 3, pp. 113-117). IEEE. 10.1109/CISP.2008.609

64. da Silva LG, da Silva Monteiro WRS, de Aguiar Moreira TM et al. Fractal dimension analysis as an easy computational approach to improve breast cancer histopathological diagnosis. Appl. Microsc. 2021;51(1):1–9. 10.1186/s42649-021-00055-w

65. Weigelt B, Horlings HM, Kreike B et al. Refinement of breast cancer classification by molecular characterization of histological special types. J. Pathol. 2008;216(2):141–150. 10.1002/path.2407

