## Supplemental Figure S1 for "Classifying breast cancer and fibroadenoma tissue biopsies from paraffined stain-free slides by fractal biomarkers in Fourier Ptychographic Microscopy"

[c] IRCCS SYNLAB SDN, Via E. Gianturco 113, Napoli, 80143, Italy.

[d] Clinica Villa Fiorita, Via Filippo Saporito 24, 81031 Aversa, Caserta, Italy.

[e] Pathological Anatomy Service, Casa di Cura Maria Rosaria, Via Colle San Bartolomeo 50, 80045 Pompei, Napoli, Italy.

*

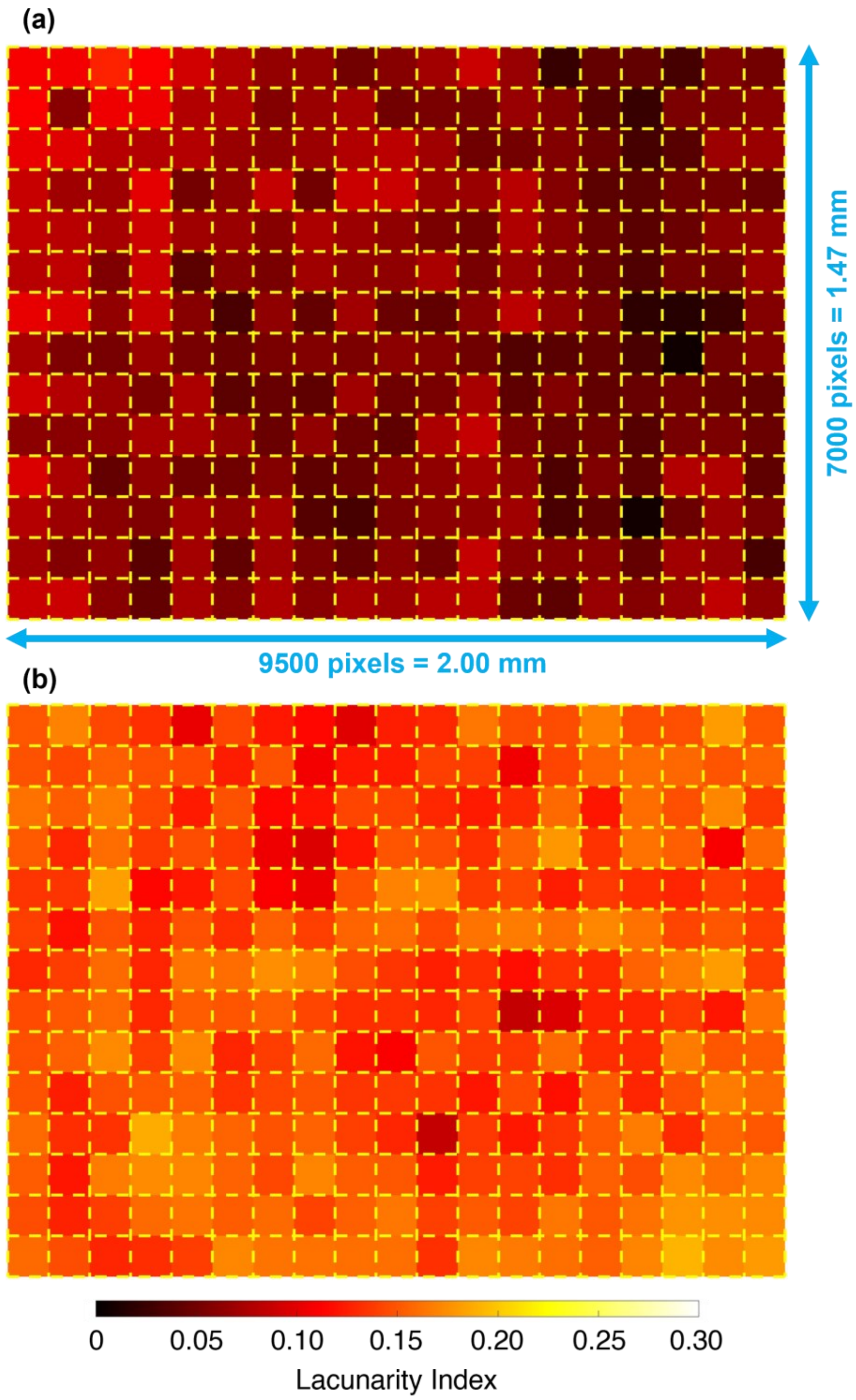


**Figure S1. Lacunarity heat map made of the lacunarity index values related to each 500x500 patch dividing the full 7000x9500 FPM FOV. (a)** Fibroadenoma tissue slide corresponding to that in Figs. 2(a,b). **(b)** Breast cancer tissue slide corresponding to that in Figs. 2(c,d).
